# Co-creating data science solutions for maternal and child health decision-making in tribal primary health centres: an action research using the Three Co’s Framework

**DOI:** 10.64898/2026.03.29.26349643

**Authors:** Arun Mitra, Gurukartick Jayaraman, Bhavana Ondopu, Santosh Kumar Malisetty, Raghu Niranjan, Shaheen Shaik, Biju Soman, Rakhal Gaitonde, Tarun Bhatnagar, Engelbert Niehaus, K.S Sajinkumar, Adrija Roy

**Affiliations:** Department of Community and Family Medicine, AIIMS Bibinagar, Hyderabad, Telangana, India; Achutha Menon Centre for Health Science Studies, Sree Chitra Tirunal Institute for Medical Sciences and Technology, Trivandrum, Kerala, India; Government Mohan Kumaramangalam Medical College, Salem, Tamil Nadu, India; Medical Officer, Primary Health Centre, Gangavaram, ASR District, Andhra Pradesh, India; Medical Officer, Primary Health Centre, Boduluru, ASR District, Andhra Pradesh, India; Medical Officer, Primary Health Centre, Vadapalli, ASR District, Andhra Pradesh, India; Achutha Menon Centre for Health Science Studies, Sree Chitra Tirunal Institute for Medical Sciences and Technology, Trivandrum, Kerala, India; ICMR - National Institute of Epidemiology, Chennai, Tamil Nadu, India; Rheinland-Pfalzische Technische Universitat Kaiserslautern-Landau, Landau, Germany; University of Kerala, Trivandrum, Kerala, India; RVM Institute of Medical Sciences and Research Centre, Siddipet, Telangana, India

## Abstract

**Background:** Digital health tools are increasingly promoted for strengthening health information systems in low- and middle-income countries, yet routine maternal and child health (MCH) data in tribal primary health centres (PHCs) in India remains underutilised for local decision-making. Top-down digital tools often fail in low-resource settings because they are designed without meaningful input from end-users. Co-creation approaches for digital health in tribal and indigenous settings are largely unexplored.

**Methods:** We conducted an action research study in three tribal PHCs under the Integrated Tribal Development Agency (ITDA), Rampachodavaram, Andhra Pradesh, India. We applied the Three Co’s Framework (Co-Define, Co-Design, Co-Refine) to co-create data science solutions for MCH decision-making with five medical officers, 24 auxiliary nurse midwives, and 36 accredited social health activists across two action research cycles (August 2023 to August 2024). Co-creation involved collaborative indicator definition, data modelling, data quality validation, health facility catchment area construction, spatial analysis, and interactive dashboard development. Keller’s Data Science Framework was employed using R to structure the analytical pipeline, and Data.org’s Data Maturity Assessment (DMA) was used to assess organisational data maturity pre- and post-intervention.

**Findings:** During Co-Define, co-creators identified a fundamental mismatch between system outputs (aggregate statistics for upward reporting) and their operational need for individual-level, geographically disaggregated, prospective information. Co-Design produced five interconnected data science solutions: (1) 42 co-defined MCH indicators grounded in clinical workflows; (2) a data model linking individuals, health services, providers, and facilities; (3) a data quality framework using the pointblank R package; (4) health facility catchment area boundaries constructed from scratch using medical officers’ local knowledge, enabling spatial analysis that revealed significant clustering of ANC coverage and anaemia prevalence; and (5) an R Shiny dashboard integrating these solutions into an offline-capable interface with lifecycle-organised views and village-level navigation. The DMA showed moderate improvement in organisational data maturity from 5.04 to 5.75 out of 10, with the largest gain in Analysis (+1.90). Co-Refine continued beyond the formal study period, with two transferred medical officers maintaining analytical engagement from new postings.

**Interpretation:** The Three Co’s Framework, combined with a data science approach, provided a structured yet flexible method for co-creating locally relevant data science solutions in a tribal setting. The framework’s explicit separation of problem definition from solution design was particularly valuable in a context where “the problem” is typically defined externally. Co-creation in tribal digital health settings is feasible and produces solutions that address locally articulated needs.

**Funding:** DST-SEED Fellowship, Government of India.

**0.1 Research in Context:** *Evidence before this study:* Co-creation and co-design approaches for digital health tools have growing evidence bases in high-income settings, but applications in low- and middle-income countries remain limited. The Three Co’s Framework was developed and applied in UK welfare contexts but has not been tested in LMIC health settings. India’s tribal populations face persistent MCH disparities, and existing digital health tools serving tribal PHCs were designed for upward reporting rather than local decision support. No published study has applied a structured co-creation framework combined with a data science approach to develop digital health tools with tribal health workers in India.

*Added value of this study:* This study provides the first application of the Three Co’s Framework in a LMIC tribal health setting. It demonstrates that structured co-creation combined with reproducible data science methods (data modelling, quality validation, spatial analysis) is feasible even under severe infrastructure constraints. The study shows that co-creators, when given genuine participatory space, reframed the problem from “how to use existing tools better” to “why existing tools do not serve local needs,” leading to fundamentally different design decisions than a top-down process would have produced.

*Implications of all the available evidence:* Co-creation frameworks developed in high-income settings can be productively adapted for LMIC tribal health contexts when combined with insider researcher positionality and flexible engagement modalities. India’s digital health strategy, including the Ayushman Bharat Digital Mission, should mandate co-creation processes for tools intended for underserved populations. The Three Co’s Framework’s explicit separation of Co-Define from Co-Design offers particular value in settings where end-users have historically been excluded from problem definition.

## 1 Introduction

Digital health dashboards are increasingly promoted as tools for strengthening local health system management in low- and middle-income countries (1,2). India’s public health infrastructure generates substantial maternal and child health data through the Reproductive and Child Health (RCH) Portal, the Health Management Information System (HMIS), and associated digital platforms. Yet at the primary health centre level, particularly in tribal areas, this data rarely informs local clinical or programmatic decisions.

This gap between data availability and data utility is well documented in the routine health information systems literature (3,4). What is less well understood is how to close it. Top-down digital health solutions, designed by developers distant from implementation contexts, frequently fail because they embed assumptions about connectivity, digital literacy, and workflow that do not match ground realities (5,6). The design-reality gap is particularly wide in tribal settings, where geographic isolation, seasonal connectivity disruptions, population mobility, and distinctive governance arrangements create conditions that generic tools cannot accommodate.

Co-creation, where end-users participate as collaborators in defining problems, designing solutions, and refining outputs, offers a pathway to contextually appropriate digital health tools. The evidence base for co-design in health is growing (7–9), but applications in tribal or indigenous digital health contexts in LMICs remain scarce (10). Most co-design studies in health originate from high-income settings and involve participants with substantially different resource contexts and digital fluency than tribal health workers in rural India.

The Three Co’s Framework, proposed by Pearce and Magee, provides a structured approach to co-creation organised around three phases: Co-Define (collaboratively identify and frame the problem), Co-Design (collaboratively develop the solution), and Co-Refine (test, iterate, and improve through field use) (11). Unlike the Double Diamond model, the Three Co’s explicitly foregrounds problem definition as a collaborative act and embeds iterative refinement as a distinct phase rather than an afterthought. The framework was developed through action research in UK welfare contexts but has not been applied in LMIC health settings.

This study is the third in a series from a data science for health project in tribal Andhra Pradesh. A companion study mapped the local MCH data ecosystem, identifying 28 data sources of which only 32% were accessible to medical officers (Mitra et al., under review). A second companion study documented the qualitative experience of data use, revealing structural constraints and complementary information practices that local decision-makers constructed to bridge the gap (Mitra et al., under review). The present study reports the action phase: co-creating data science solutions for MCH decision-making with frontline health workers using the Three Co’s Framework. We describe the co-creation process, the suite of solutions produced (indicator definitions, data model, quality framework, spatial resources, and interactive dashboard), and changes in organisational data maturity.

## 2 Methods

### 2.1 Study design

This study was conducted as part of an action research project following two iterative cycles (12). The Three Co’s Framework (11) structured the co-creation process across these cycles. The first cycle (diagnosing and action planning) corresponded to the Co-Define phase. The second cycle (action taking, evaluating, and specifying learning) encompassed Co-Design and Co-Refine (13). The data science approach followed the framework proposed by Keller et al., which organises data work through data discovery, data preparation, data quality assessment, and analysis (14).

### 2.2 Setting

The study was conducted in three PHCs under ITDA Rampachodavaram, Alluri Sitharama Raju (ASR) District, Andhra Pradesh. ASR District was constituted in April 2022 by carving the tribal-majority portions of East Godavari and Visakhapatnam districts, creating a predominantly tribal district with over 70% Scheduled Tribe population. The ITDA Rampachodavaram, established under the Fifth Schedule of the Indian Constitution, administers seven mandals covering 4,433 km^2^ of hilly, forested terrain with 818 habitations and 61,013 households.

The three study PHCs represent variation in geography, connectivity, and tribal composition (Table 1). PHC Boduluru (Maredumilli mandal) is the most remote site, with 81.6% forest cover, 93.3% ST population, and the highest PVTG (Particularly Vulnerable Tribal Group) concentration at 39.8% of the ST population. PHC Gangavaram (Gangavaram mandal) has moderate connectivity and 67.2% ST population. PHC Vadapalli (Rampachodavaram mandal) has 79.3% ST population with limited connectivity. Each PHC has one to two medical officers, two to three staff nurses, and coordinates with four to eight sub-centres staffed by ANMs and ASHAs. Mobile network coverage is inconsistent across all three sites, with seasonal disruptions during monsoon months (June to September).

**Table 1:** Study site characteristics. Mandal-level data from ITDA Rampachodavaram administrative records and Census 2011.

| Characteristic | Boduluru | Gangavaram | Vadapalli |
| --- | --- | --- | --- |
| Mandal | Maredumilli | Gangavaram | Rampachodavaram |
| Mandal area (km<sup>2</sup>) | 913.96 | 612.71 | 610.30 |
| Forest cover (%) | 81.6 | 69.4 | 65.1 |
| Village Secretariats (n) | 8 | 10 | 16 |
| Habitations (n) | 113 | 79 | 102 |
| Scheduled Tribe population (%) | 93.3 | 67.2 | 79.3 |
| PVTG population (% of ST) | 39.8 | 1.3 | 2.4 |
| Connectivity | Limited | Moderate | Limited |

### 2.3 Co-creators

Co-creators included five medical officers across the three PHCs, 24 ANMs, and 36 ASHAs. Medical officers served as the primary co-design partners given their role as local MCH decision-makers. ANMs and ASHAs participated through weekly convergence meetings (Thursdays at all three PHCs) and contributed field-level insights on data workflows and beneficiary tracking. Additional key informants included the Additional District Medical and Health Officer and two data entry operators at the ITDA level, including the Multipurpose Health Data Entry Operator (MPHDEO). Following Pearce and Magee’s recommendation, we refer to participants as co-creators throughout.

### 2.4 The Three Co’s process

#### 2.4.1 Co-Define (August 2023 to January 2024)

Co-Define corresponded to the diagnostic phase of the action research project. Activities included systematic mapping of the local MCH data ecosystem (identifying 28 data sources across healthcare and social determinant domains), key informant interviews with medical officers and data entry operators (n=8), participant observation at weekly convergence meetings, and document review of registers and reporting formats. The researcher shared preliminary analyses of RCH Portal data with medical officers, asked interpretive questions, and gathered contextual insights through structured dialogue.

A critical output of Co-Define was the collaborative identification of MCH indicators that medical officers needed for daily decision-making. Through iterative discussions, co-creators specified 42 indicators across four domains: maternal health (registration, ANC visits, haemoglobin levels, high-risk flagging, tetanus toxoid coverage), child health (birth weight, immunisation status by antigen, immunisation timeliness), service delivery (delivery type, institutional delivery, PNC visits), and geographic coverage (village-level and sub-centre-level aggregations). These indicator definitions were grounded in clinical workflows rather than programme reporting requirements. For example, co-creators defined “ANC pending” not by the system’s rigid cutoff dates but by the clinically relevant interval since the last visit, addressing a long-standing frustration with the RCH Portal’s inflexible logic.

The baseline Data Maturity Assessment (DMA), a structured tool from data.org measuring organisational data maturity across Purpose, Practice, and People dimensions, was administered to all five medical officers in August 2023.

#### 2.4.2 Co-Design (February to August 2024)

Co-Design translated the diagnostic findings into a functional dashboard through three workstreams: data modelling, data quality validation, and interface prototyping.

### Data modelling

The first technical step was constructing a data model that reflected the MCH service delivery context. Through iterative discussions with medical officers, an entity-relationship model was developed linking five core entities: INDIVIDUAL (identified by RCHID, representing mothers and children), HEALTH-SERVICE (service type, date, clinical details), HEALTH-PROVIDER (ANM, ASHA), HEALTH-FACILITY (PHC, sub-centre), and HEALTH-SERVICE-LOCATION (village, distances to facilities). This model structured the data pipeline and determined how the dashboard would enable drill-down from PHC-level summaries to individual beneficiary records.

### Data quality validation

Raw RCH Portal data contained systematic quality issues that had to be addressed before any solution could provide reliable information (15–17). Using the pointblank package in R, two validation agents were constructed collaboratively with medical officers. The maternal health validation agent checked RCHID format (128 prefix followed by 9 digits), date sequencing (LMP before registration, registration before delivery), haemoglobin values within physiological range, and mother age between 12 and 55 years. The child health validation agent checked birth weight within 0.5 to 6.0 kg, vaccine date sequencing relative to date of birth, and immunisation schedule adherence (BCG and OPV-0 within 15 days of birth; Pentavalent-1 at 6 weeks; Measles at 9 months). Medical officers reviewed validation outputs and flagged rules that needed adjustment based on local clinical experience, such as relaxing the birth weight lower bound for the high prevalence of low birth weight in PVTG communities.

### Interface prototyping

Traditional co-design workshops proved infeasible due to medical officers’ clinical workload and unreliable internet. The process adapted to a distributed model: short design conversations during field visits, screen-sharing demonstrations, and asynchronous WhatsApp feedback. A pivotal moment occurred on 26 February 2024, when the researcher invited a medical officer at PHC Boduluru to sketch his ideal dashboard on paper. The resulting sketch became the foundational prototype.

Through three to four iterations per PHC, five design principles crystallised from co-creator input:

1. **Actionable over informative.** Every displayed element should support a specific decision. Pending action lists took priority over completed counts.
2. **Names over numbers.** Individual beneficiary information rather than aggregate statistics.
3. **Drill-down by default.** Progressive navigation from PHC to sub-centre to village to individual, matching how medical officers conceptualise their service area.
4. **Familiar over novel.** Interface patterns aligned with WhatsApp and spreadsheets, tools co-creators already used daily.
5. **Supplement, not replace.** The dashboard complemented existing workflows rather than requiring new routines.

**Data pipeline and spatial analysis.** The data pipeline was built using the targets framework in R (18), processing raw RCH Portal extracts through eight sequential steps: data loading and cleaning, derived variable construction, data quality validation, variable documentation, data linkage, data integration, exploratory data analysis (EDA), and exploratory spatial data analysis (ESDA). The pipeline operated offline through daily synchronisation using GitHub Actions.

A notable example of co-creation driving technical work involved spatial data. Medical officers expressed a need for map-based views of MCH indicators, but no health facility catchment area (HFCA) boundaries existed for the ITDA (19,20). Administrative shapefiles did not align with PHC service areas. Through iterative back-and-forth with medical officers, who provided local knowledge of village-to-sub-centre assignments, the researcher constructed HFCA boundaries from scratch. ESDA using these co-created boundaries revealed significant spatial clustering of both ANC coverage (Moran’s I=0.28, p<0.001) and anaemia prevalence (Moran’s I=0.28, p<0.001) across sub-centre catchment areas, confirming that the geographic targeting medical officers had requested was analytically justified.

The resulting R Shiny dashboard (21,22) organised MCH data along the lifecycle continuum (pregnancy registration through child immunisation) with village-level navigation at each level. The approach built on prior work by the research team in developing data science decision support tools for public health (23,24).

#### 2.4.3 Co-Refine (August 2024 to ongoing)

Co-Refine adapted to a significant contextual challenge: medical officer transfers. Two of the five original co-creators were transferred to other districts during the study period. Rather than ending their participation, both maintained engagement via WhatsApp, offering feedback on dashboard iterations from their new postings.

A structured co-refine session on 15 December 2024, when the PhD supervisor visited PHC Gangavaram, provided formal review with the medical officer still in post. The visit combined dashboard demonstration with structured feedback, and the supervisor’s presence signalled institutional commitment to the co-creation process.

The post-intervention DMA was administered in August 2024, twelve months after baseline.

### 2.5 Researcher positionality and ethics

The lead researcher (AM) belongs to the tribal community in the study area. For the co-creation process, this positionality was consequential in two ways. First, it enabled the iterative, trust-dependent engagement model that the Three Co’s process required: short, informal design conversations during field visits, asynchronous WhatsApp exchanges, and candid feedback that would have been difficult to achieve with an outsider researcher. Second, it introduced a productive tension between the researcher’s analytical perspective (trained in data science and epidemiology) and the co-creators’ operational perspective (grounded in clinical workflows and local knowledge). This tension generated design decisions that neither perspective would have reached alone. Reflexive strategies included regular supervisor debriefs, independent coding by a second analyst, and systematic attention to moments where the researcher’s assumptions diverged from co-creator priorities.

The study received ethical approval from the Institutional Ethics Committee, Sree Chitra Tirunal Institute for Medical Sciences and Technology (Protocol No. SCT/IEC/2047/MAY/2023), and followed ITDA community research protocols. The CONSIDER statement for research involving Indigenous peoples informed the ethical framework (25). All participants provided written informed consent. Transferred medical officers provided renewed consent for continued participation via WhatsApp.

## 3 Results

### 3.1 Co-Define: reframing the problem

The diagnostic phase revealed that medical officers did not lack data; they lacked access to their own data in actionable forms. The Co-Define process mapped 28 distinct MCH data sources across the three PHCs (Mitra et al., under review). Despite this abundance, medical officers consistently described being unable to answer basic operational questions: which beneficiaries in which villages were overdue for which services.

A critical reframing occurred during Co-Define. The researcher initially approached the problem as one of data analysis capacity. Co-creators redirected this framing toward data access and system architecture. Formal health information systems channelled data toward district and state supervisory functions without providing facility-level decision support in return (5). The problem was structural, not individual. This reframing shaped the subsequent design phase: the dashboard needed to bridge an access gap, not fill a skills gap.

The co-defined indicator set reflected this reframing. Rather than the aggregate statistics that formal systems produced, co-creators specified indicators tied to individual beneficiaries and geographic units. For maternal health, this meant tracking each woman’s ANC visit count against her gestational age, categorising haemoglobin levels by clinical severity (severe <7 g/dL, moderate 7-10 g/dL, normal >10 g/dL), and flagging high-risk pregnancies for follow-up. For child health, this meant tracking each child’s immunisation status against the national schedule with specific windows (BCG within 15 days, Pentavalent-1 at 6 weeks, Measles at 9 months) and highlighting overdue vaccinations by village. These indicator definitions became the functional requirements for the dashboard.

The baseline DMA confirmed this diagnosis quantitatively (Table 2). The overall score of 5.04 out of 10 (“Data Guided” level) revealed a telling pattern: Purpose scored highest (6.58), driven by Application (8.66), indicating that medical officers valued and wanted to use data. But Practice scored lowest (4.24), with Security at 2.12 and Infrastructure at 4.70, reflecting structural barriers. Within Purpose, the gap between Application (8.66) and Analysis (4.78) became the central design target: medical officers valued data but lacked tools to act on it.

**Table 2:** Data Maturity Assessment scores pre- and post-intervention (mean across three PHCs, n=5 medical officers). Baseline scores verified from individual DMA result PDFs. Post-intervention scores from analysis pipeline output. Scale: 0-10.

| Category | Subcategory | Baseline | Post | Change |
| --- | --- | --- | --- | --- |
| **Purpose** |  | 6.58 | 7.40 | +0.82 |
|  | Application | 8.66 | 9.13 | +0.47 |
|  | Analysis | 4.78 | 6.68 | +1.90 |
|  | Strategy | 5.92 | 7.05 | +1.13 |
| **Practice** |  | 4.24 | 4.17 | -0.07 |
|  | Quality | 4.42 | 5.23 | +0.81 |
|  | Security | 2.12 | 1.80 | -0.32 |
|  | Responsible Use | 6.18 | 6.00 | -0.18 |
|  | Infrastructure | 4.70 | 4.57 | -0.13 |
| **People** |  | 5.00 | 5.77 | +0.77 |
|  | Leadership | 5.52 | 6.37 | +0.85 |
|  | Talent | 3.52 | 4.50 | +0.98 |
|  | Culture | 6.02 | 6.70 | +0.68 |
| **Overall** |  | **5.04** | **5.75** | **+0.71** |

**Table 3:** Co-creator engagement in the Three Co’s process.

| Co-creator | PHC | Role in co-creation | Engagement | Status (2026) |
| --- | --- | --- | --- | --- |
| MO-1 | Bodu-luru | Primary co-designer; foundational sketch; indicator definitions | High | Transferred; ongoing WhatsApp |
| MO-2 | Vada-palli | Consistent feedback; data quality rule refinement | Good | Transferred; ongoing |
| MO-3 | Gan-gavaram | Supervisor visit review; sustained field engagement | Moderate | In post; most active |
| MO-4 | Bodu-luru | WhatsApp feedback | Low | In post |
| MO-5 | Vada-<br>palli | Minimal visible engagement | Low | In post |

### 3.2 Co-Design: data science solutions

The Co-Design phase produced five interconnected data science solutions: co-defined MCH indicators, a data model, a data quality framework, health facility catchment area boundaries with spatial analysis, and an interactive dashboard.

The entity-relationship model (Figure 1) structured data around five entities reflecting the MCH service delivery context. The INDIVIDUAL entity, keyed on the RCHID (Reproductive and Child Health Identifier), linked mothers and children to HEALTH-SERVICE events, which in turn connected to HEALTH-PROVIDER (ANM or ASHA), HEALTH-FACILITY (PHC or sub-centre), and HEALTH-SERVICE-LOCATION (village, with distances to nearest facilities). This model enabled the drill-down navigation that co-creators had specified as a core requirement.

**Figure 1:**
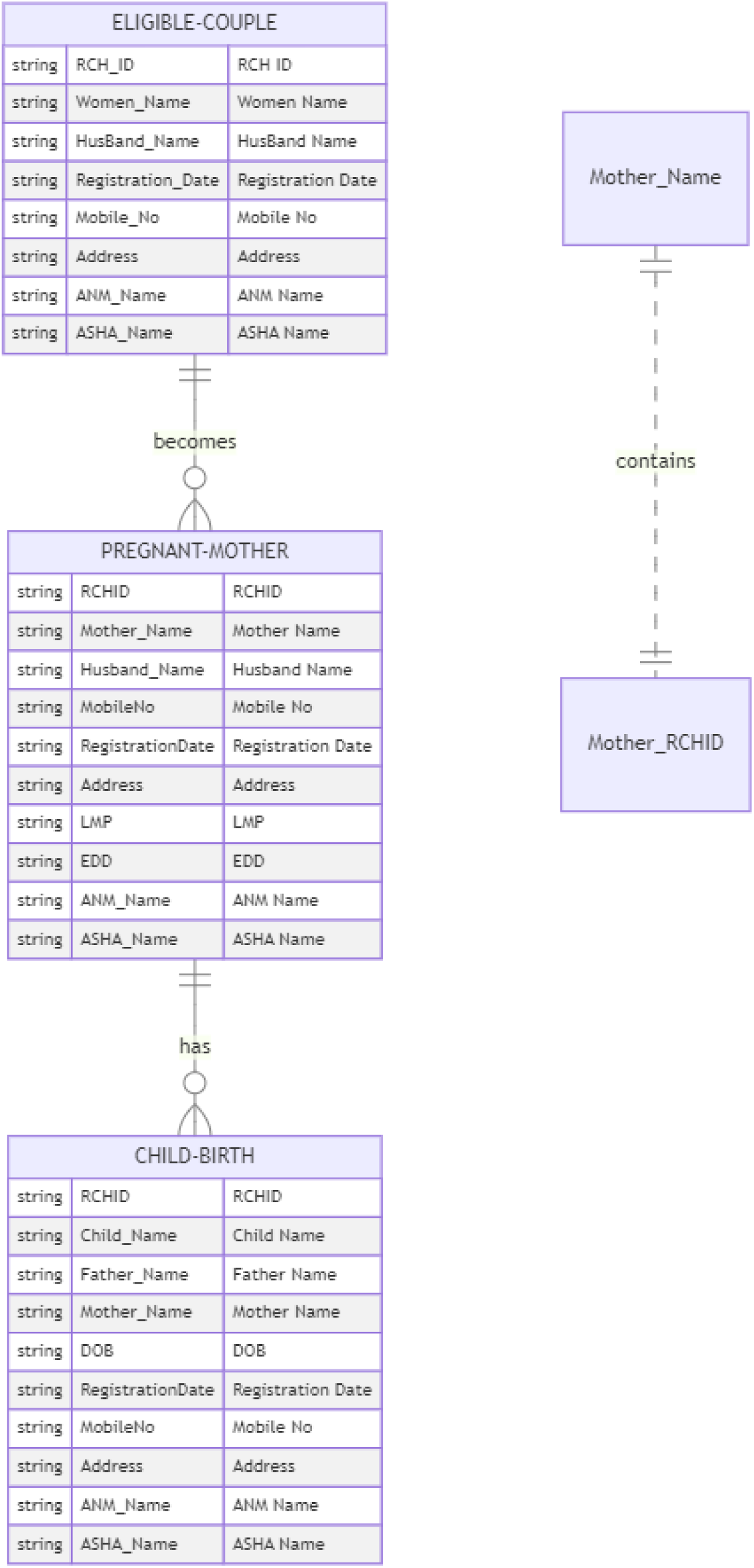
Entity-relationship diagram of the MCH data model co-developed with medical officers. Five linked entities (Individual, Health-Service, ^7^Health-Provider, Health-Facility, Health-Service-Location) structure the data pipeline and enable drill-down navigation from facility to individual level.

**Figure 2:**
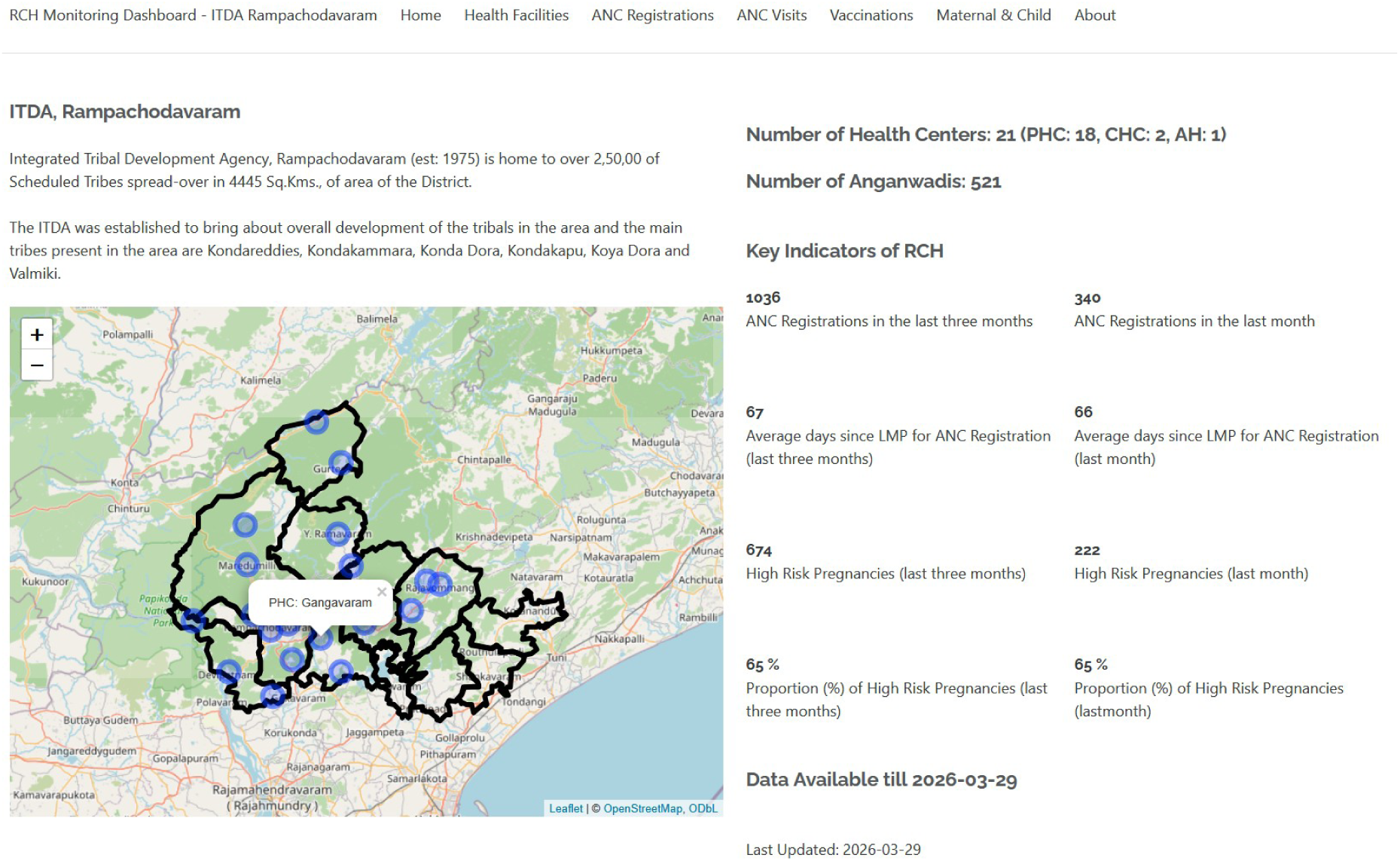
The co-created RCH Monitoring Dashboard. The landing view shows the ITDA-level summary with an interactive map of health centres, key MCH indicators (ANC registrations, high-risk pregnancies), and navigation to facility-level drill-down views.

**Figure 3:**
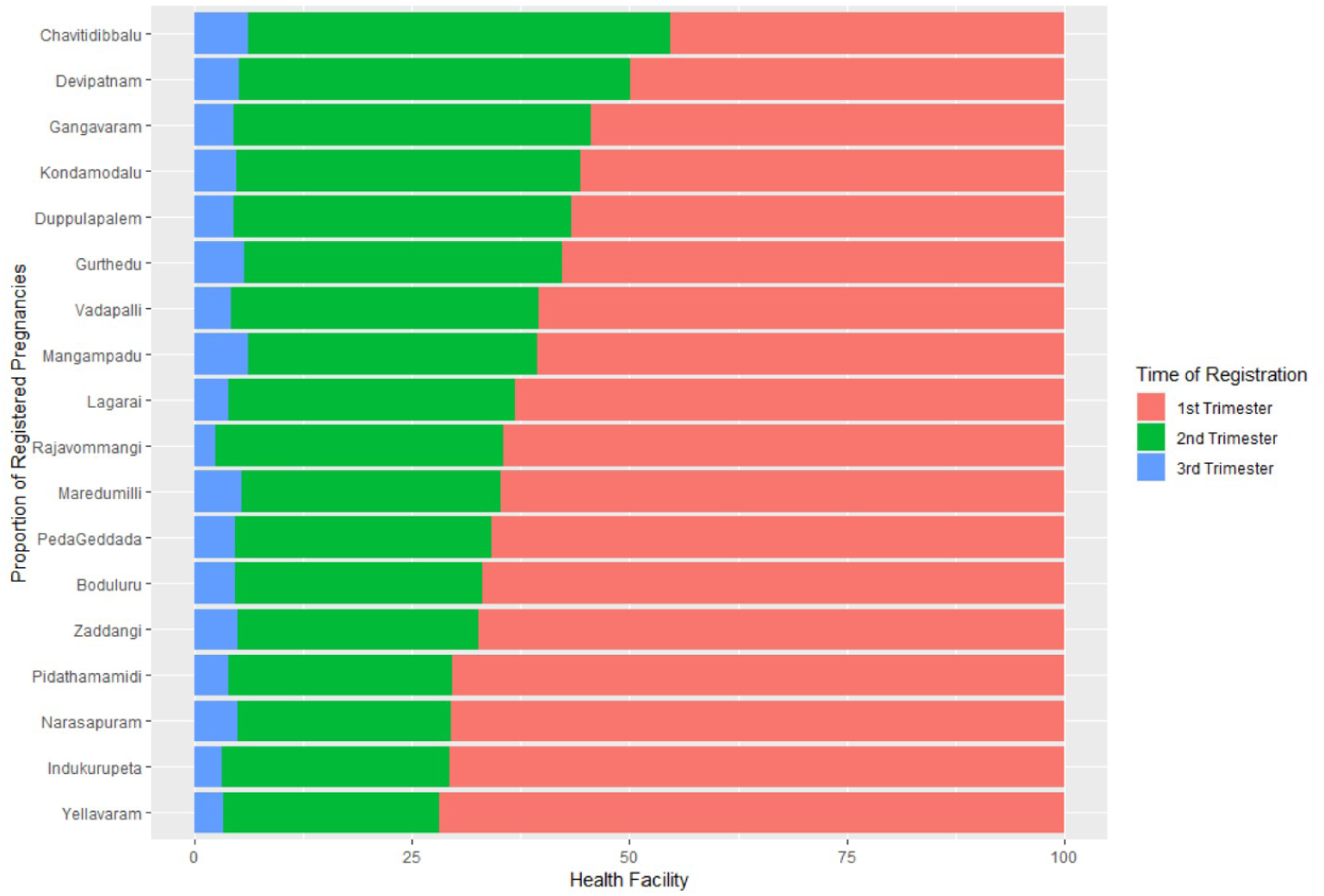
Facility-level drill-down showing ANC registration by trimester across all PHCs in ITDA Rampachodavaram. This view directly implements the “names over numbers” and “drill-down by default” design principles, enabling medical officers to compare facilities and identify those with late registrations.

**Figure 4:**
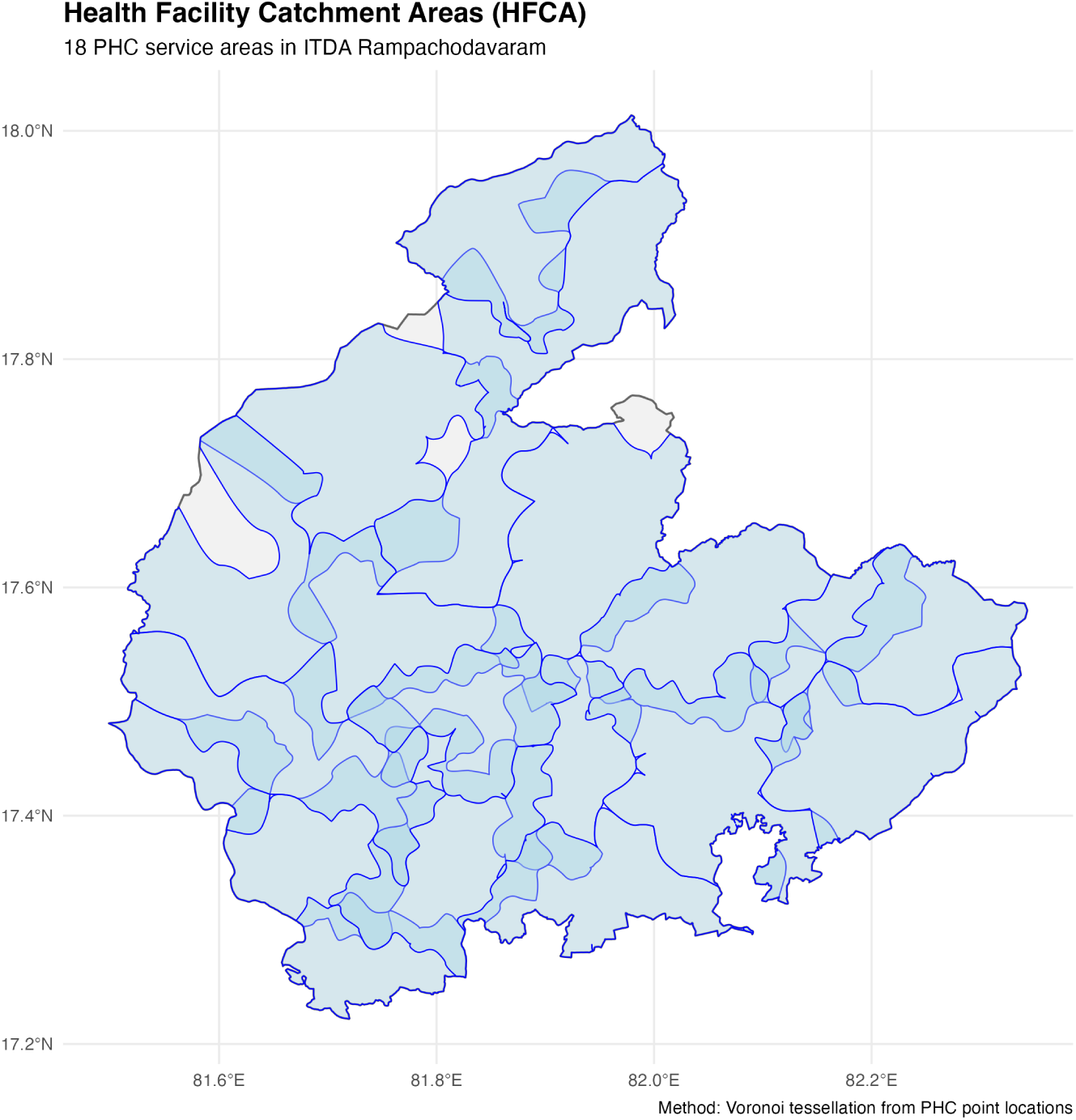
Health facility catchment area (HFCA) boundaries co-constructed with medical officers using Voronoi tessellation from PHC point locations. These boundaries did not previously exist and were created through iterative exchange between the researcher’s spatial analysis and medical officers’ local knowledge of village-to-sub-centre assignments. Left: 18 PHC-level catchment areas. Right: 96 Village Secretariat (sub-centre) catchment areas.

**Figure 5:**
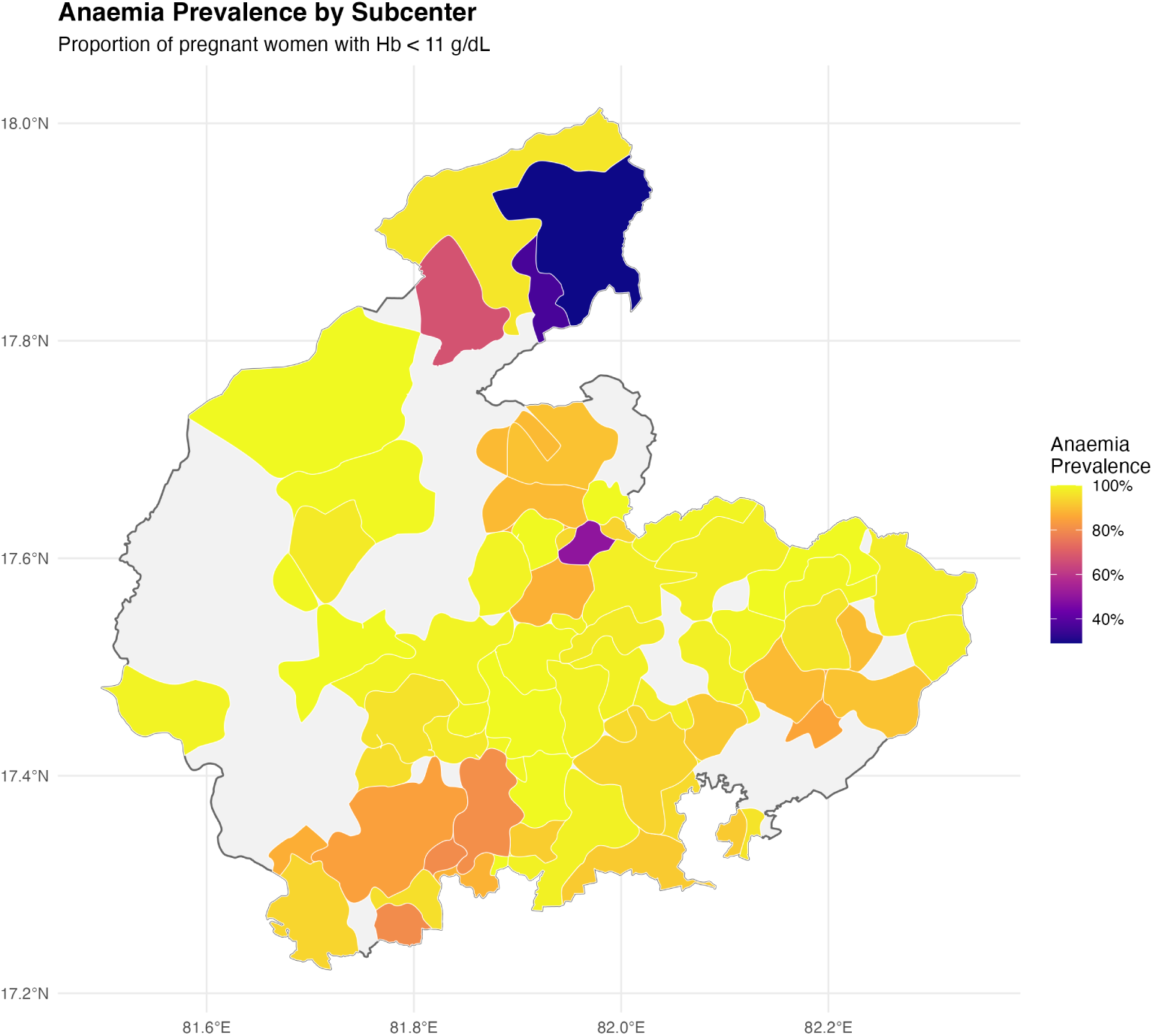
Anaemia prevalence by sub-centre catchment area (left) and Local Indicators of Spatial Association (LISA) clusters (right). Moran’s I = 0.28 (p<0.001) confirmed significant spatial clustering, validating the geographic targeting that medical officers had requested.

**Figure 6:**
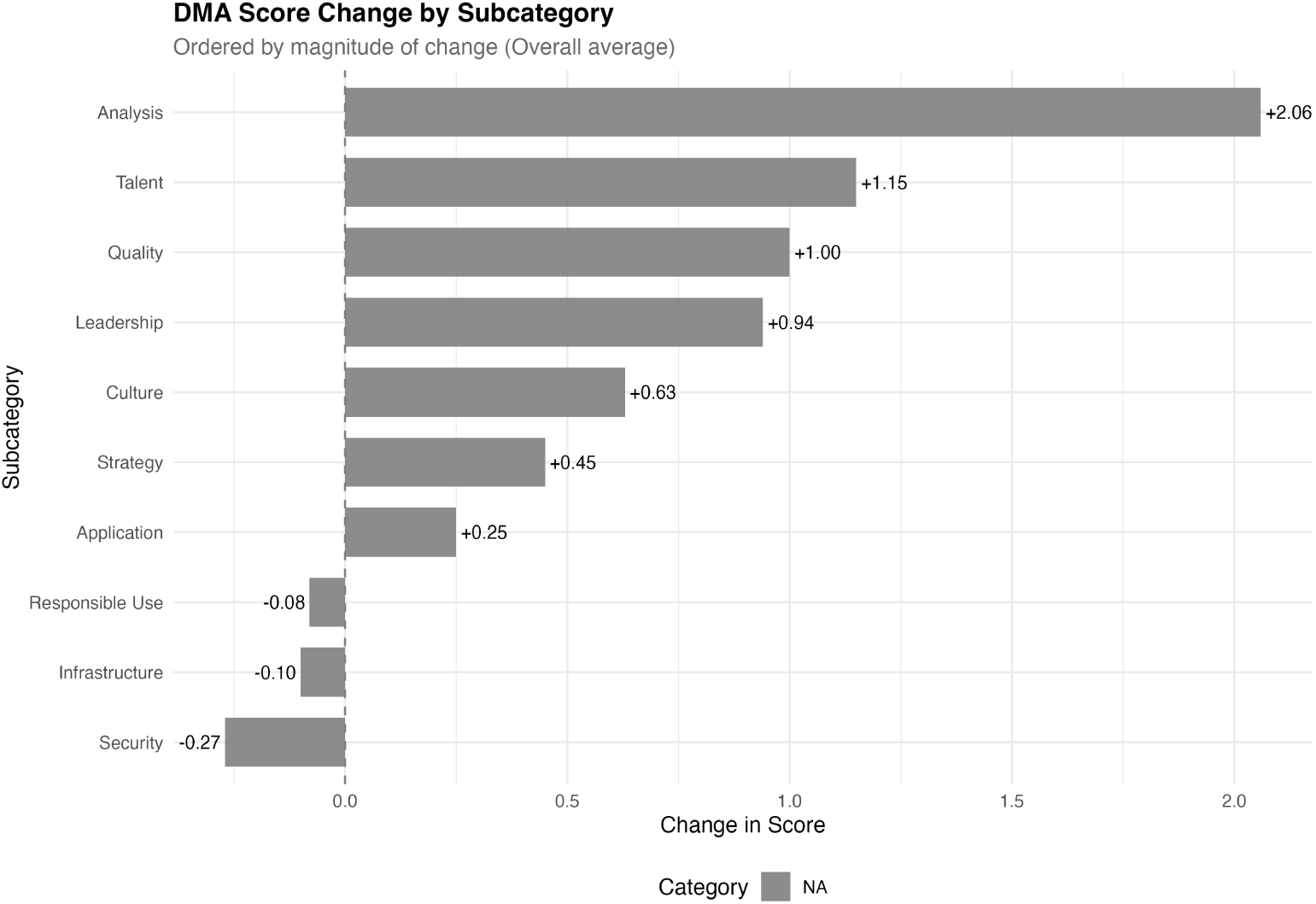
DMA score change by subcategory, ordered by magnitude. Analysis (+2.06) and Talent (+1.15) showed the largest gains. Practice-level subcategories (Security, Infrastructure, Responsible Use) showed no improvement or slight decline, reflecting structural constraints beyond the scope of the co-created solutions.

The data quality framework addressed systematic issues in RCH Portal data. Validation agents built with the pointblank package applied tiered checks: critical validations (RCHID format, null checks), important validations (date sequencing, physiological ranges), and advisory validations (completeness thresholds, duplicate detection). Medical officers reviewed validation reports and refined rules iteratively. For instance, the initial haemoglobin range validation (7-16 g/dL) was broadened after co-creators noted that severely anaemic women with Hb below 7 g/dL were being excluded as data errors rather than flagged as clinical priorities.

The interactive dashboard integrated these solutions into a single offline-capable R Shiny interface. The underlying data pipeline, built with the targets framework, processed raw RCH Portal extracts through cleaning, deduplication, entity linkage, indicator calculation, and spatial aggregation. The interface organised MCH data along the lifecycle continuum with geographic drill-down at each level, directly implementing the five design principles articulated during Co-Design. The dashboard was thus not the sole output of co-creation but the integration layer that brought together the indicator definitions, data model, quality framework, and spatial resources into a usable tool.

### 3.3 Co-Refine: sustained engagement and organisational change

Two transferred medical officers maintained engagement via WhatsApp from new postings, offering feedback on dashboard iterations. In a WhatsApp exchange in June 2025, one transferred medical officer independently analysed temporal trends in MCH staffing data, contextualising a data visualisation with local knowledge. This unprompted analytical engagement, more than a year after transfer, suggests that capacities developed through co-creation were internalised rather than remaining tied to the study setting. This resonates with normalisation process theory’s concept of cognitive participation: once individuals invest in understanding and valuing a new practice, that investment becomes part of their professional identity (26).

The post-intervention DMA, administered twelve months after baseline, showed moderate improvement in organisational data maturity (Table 2). The overall score improved from 5.04 to 5.75 (+0.71). The pattern was instructive: Purpose improved (+0.62), driven by Analysis (+2.06), the dimension most directly targeted by the dashboard. People improved more substantially (+0.84), with Talent (+1.15) and Leadership (+0.94) suggesting that the co-creation process itself built capacity beyond the dashboard artefact. Practice remained unchanged (4.24 to 4.17): the dashboard could not address infrastructure deficits, connectivity constraints, or data security weaknesses that require system-level reform. These results should be interpreted cautiously given the small sample (n=5), absence of a control group, and the possibility that observed changes reflect broader influences beyond the intervention.

## 4 Discussion

### 4.1 The value of collaborative problem definition

The Co-Define phase proved more consequential than anticipated. When given genuine space to define the problem, co-creators reframed it from a skills issue to a structural design issue. This reframing is consistent with the broader co-design literature (7) but takes on particular significance in tribal health settings, where problems are typically defined by external actors (10). The Three Co’s Framework’s explicit separation of problem definition from solution design created space for this reframing, an architectural feature that distinguishes it from the Double Diamond model.

The collaborative indicator definition process was central to this phase. Co-creators did not want more data; they wanted the existing data restructured around clinical decision points. The shift from system-defined “pending” (based on rigid reporting cutoffs) to clinically defined “pending” (based on gestational age and visit intervals) illustrates how co-creation can produce analytically different outputs than top-down design.

### 4.2 Data science as a co-creation medium

The data science pipeline served as a medium for co-creation rather than merely a technical backend. Data quality validation with pointblank became a collaborative activity (27): medical officers reviewed validation reports and refined rules based on clinical knowledge that the researcher could not have applied alone. The HFCA boundary construction required iterative exchange between the researcher’s spatial analysis capability and the medical officers’ local knowledge of village-to-sub-centre assignments, extending earlier work on reproducible spatiotemporal frameworks for health data in India (28). The ESDA outputs (spatial clustering of ANC coverage and anaemia prevalence) provided analytical evidence that the geographic targeting co-creators had intuitively requested was statistically justified.

This integration of data science within a co-creation framework extends the Three Co’s Framework by demonstrating that technical work (data modelling, quality validation, spatial analysis) can be a participatory activity rather than something done to or for end-users.

### 4.3 Adapting co-creation for resource-constrained settings

The co-creation process required substantial adaptation from its original formulation. Workshop-based engagement was impractical given medical officers’ clinical workload and unreliable internet. The distributed, episodic model that emerged (short field visit conversations, asynchronous WhatsApp exchanges, brief screen-sharing sessions) was a contextual adaptation that produced authentic engagement grounded in daily practice rather than artificial scenarios. This finding extends the Three Co’s Framework by demonstrating that its phases can be operationalised through flexible modalities rather than requiring structured workshop formats (8).

### 4.4 Organisational data maturity as a co-creation outcome

The DMA results provide modest evidence that the co-creation process contributed to improved organisational data maturity (29), particularly in Analysis and Talent dimensions. However, these results should not be overstated. The sample was small (n=5), there was no control group, and multiple concurrent factors (district reorganisation, staff transfers, evolving digital health policies) may have influenced the scores. The flat Practice score (4.24 to 4.17) is arguably the most important finding: it demonstrates that co-created data science solutions, however contextually grounded, cannot address infrastructure deficits, connectivity constraints, and data security weaknesses that require system-level reform. The DMA is useful as a structured description of change, not as evidence of causal impact.

### 4.5 Limitations

The study involved three facilities within a single ITDA jurisdiction, offering contextual depth but limiting generalisability to other tribal or non-tribal settings. The Co-Refine phase is ongoing, and long-term sustainability is not yet demonstrated. Differential engagement across co-creators means the “co” in co-creation was more fully realised with some participants than others. The DMA pre/post comparison lacks a control group, and the small sample precludes statistical inference.

The insider researcher positionality, while enabling trust, may have introduced assumptions that external researchers would not share; this was managed through independent coding, supervisor debriefs, and systematic attention to disconfirming evidence.

### 4.6 Implications

For practice, the study demonstrates that co-creation combined with data science methods is feasible in tribal health settings when engagement modalities are adapted to context. The individual solutions (indicator definitions, data quality agents, HFCA boundaries, spatial resources) are independently transferable to other tribal PHC settings using the RCH Portal. For policy, India’s Ayushman Bharat Digital Mission (30) should mandate co-creation processes for digital tools intended for underserved populations, with the Co-Define phase serving as a diagnostic before any tool is deployed. For research, comparative applications of the Three Co’s Framework across different LMIC health settings would test the framework’s transferability.

## 5 Conclusion

The Three Co’s Framework, combined with a data science approach, enabled the co-creation of a suite of MCH data science solutions that address locally articulated needs rather than externally assumed ones. The explicit separation of Co-Define from Co-Design was the framework’s most valuable feature, creating space for co-creators to reframe the problem before solutions were proposed. The five co-created solutions (indicator definitions, data model, quality framework, spatial resources, and interactive dashboard) are individually transferable to other tribal PHC settings using the RCH Portal, and collectively demonstrate that data science for health can be a participatory practice rather than a top-down technical exercise. The DMA provides modest evidence of improved organisational data maturity, though the impact on Practice-level constraints (infrastructure, security) remains bounded by structural factors beyond the scope of co-created solutions.

## Declarations

### Ethics approval and consent to participate

Approved by the Institutional Ethics Committee, Sree Chitra Tirunal Institute for Medical Sciences and Technology (Protocol No. SCT/IEC/2047/MAY/2023). The CONSIDER statement for research involving Indigenous peoples informed the ethical framework (25). Transferred medical officers provided renewed consent for continued participation.

## Consent for publication

Not applicable.

### Availability of data and materials

The dashboard source code and data pipeline are available from the corresponding author on reasonable request. The DMA instrument and scoring are available from data.org.

## Authors’ contributions

AM conceptualised the study, conducted all fieldwork, developed the data pipeline and dashboard, led the analysis, and drafted the manuscript. AM and GJ independently coded process data; discrepancies were resolved through deliberation. GJ contributed to field data collection. BO, SKM, RN, and SS, as PHC Medical Officers and co-creators, provided indicator definitions, data quality feedback, design input, and reviewed the draft. RG and BS supervised the study design and critically revised the manuscript. BS conducted a field verification visit in December 2024. TB, EN, and SK served on the doctoral advisory committee and contributed to conceptualisation, review of findings, and structuring of the draft. AR contributed to field data collection. All authors reviewed and approved the final version.

## Data Availability

All data produced in the present study are available upon reasonable request to the authors

## Acknowledgements

We thank the ANMs, ASHAs, and data entry operators at the study PHCs for their partnership as co-creators. We acknowledge the ITDA Rampachodavaram administration for facilitating access.

